# Use of focal radiotherapy boost for prostate cancer and perceived barriers toward its implementation: a survey

**DOI:** 10.1101/2023.02.01.23285345

**Authors:** Allison Y. Zhong, Asona J. Lui, Matthew S. Katz, Alejandro Berlin, Sophia C. Kamran, Amar U. Kishan, Vedang Murthy, Himanshu Nagar, Daniel Seible, Bradley J. Stish, Alison C. Tree, Tyler M. Seibert

## Abstract

**Background:** In a recent phase III randomized control trial (FLAME), delivering a focal radiotherapy (RT) boost to tumors visible on MRI was shown to improve outcomes for prostate cancer patients without increasing toxicity. The aim of this study was to assess how widely this technique is being applied in current practice as well as physicians’ perceived barriers toward its implementation.

**Methods:** An online survey assessing the use of intraprostatic focal boost was conducted in December 2022 and February 2023. The survey link was distributed to radiation oncologists worldwide via email list, group text platform, and social media.

**Results:** The survey initially collected 205 responses from various countries over a two-week period in December 2022. The survey was then reopened for one week in February 2023 to allow for more participation, leading to a total of 263 responses. The highest-represented countries were the United States (42%), Mexico (13%), and the United Kingdom (8%). The majority of participants worked at an academic medical center (52%) and considered their practice to be at least partially genitourinary (GU)-subspecialized (74%).

57% of participants reported *not* routinely using intraprostatic focal boost. Even among complete subspecialists, a substantial proportion (39%) do not routinely use focal boost. Less than half of participants in both high-income and low-to-middle-income countries were shown to routinely use focal boost. The most commonly cited barriers were concerns about registration accuracy between MRI and CT (37%), concerns about risk of additional toxicity (35%), and challenges to accessing high-quality MRI (29%).

**Conclusion:** Despite level 1 evidence from the FLAME trial, most radiation oncologists surveyed are not routinely offering focal RT boost. Adoption of this technique might be accelerated by increased access to high-quality MRI, better registration algorithms of MRI to CT simulation images, physician education on benefit-to-harm ratio, and training on contouring prostate lesions on MRI.

## Introduction

A phase III randomized controlled trial (FLAME) demonstrated that a focal radiotherapy (RT) boost to tumors visible on MRI improves outcomes for patients with intermediate- and high-risk prostate cancer^1^. Participants were randomly assigned to receive either uniform RT dose to the entire prostate (control arm) or RT to the entire prostate with a focal RT dose boost to gross disease (focal boost arm). Compared to the control arm, participants in the focal boost arm had improved disease-free survival, improved local control, and improved regional/distant metastasis-free survival^1,2^. No difference in toxicity was observed between the two groups^1^. Thus, there is level 1 evidence that a meaningful oncologic benefit can be offered patients with prostate cancer without increased side effects. This approach can be delivered on RT equipment already widely available for clinical use. Two years after first publication of the FLAME trial, we sought to learn whether patients are currently able to access this benefit.

Differential adoption of focal boost may have introduced a new healthcare disparity for patients with prostate cancer. Information about radiation dose and use of focal RT boost is not routinely or publicly available. Patients may not be aware of whether they are receiving focal boost or whether this approach was even considered for them. We decided to directly survey radiation oncologists to ask if they have adopted the focal boost approach. If some oncologists are offering focal boost and others are not, this would clearly imply a disparity in practice that has been shown to affect outcomes. We also asked survey respondents about perceived barriers to implementation of focal boost in their own practice.

## Methods

In December 2022 and February 2023, we conducted an international survey of radiation oncologists. In designing the survey, we recognized two challenges. First, we are not aware of a global list of all radiation oncologists, which would be required for a survey of the complete population or to identify a random sample of that population. Second, even if all radiation oncologists could be contacted, it is likely only a small percentage would choose to participate, making accurate generalization of results impossible. Thus, robust generalizability may not be feasible. Still, a survey with a large number of responses from a diverse group of participants can be informative about practice patterns. We opted for a pragmatic approach: a group of authors from varied practice settings (country of practice; academic or private; urban, suburban, or rural) agreed to advertise the survey through electronic media. While this approach would not allow formal generalization of results or calculation of a response rate (as the number of radiation oncologists contacted via social media is not known), we would be able to cast a wide net and obtain enough responses to meet the primary study goals: (1) determine whether a substantial group of radiation oncologists exists that has not already adopted focal boost for prostate cancer, and (2) gain some insight into perceived barriers to adoption.

The study was approved by the Institutional Review Board at UC San Diego. Participants gave consent electronically. The survey was designed to be very brief to encourage participation (no more than 10 minutes, with initial feedback suggesting typical completion in less than 3 minutes).

We advertised the survey to our respective contacts via email and social media. We also used a previously curated email list of 850 members of the American Society for Radiation Oncology (ASTRO) practicing in the New England region of the United States and a group text-message platform for members of Sociedad Mexicana de Radioterapeutas (SOMERA) with 291 users, most of which are radiation oncologists. Participants were asked to consent electronically; they self-reported as practicing radiation oncologists who treat patients with prostate cancer.

The survey included 12 questions (Supplementary Material). We asked participants whether they use focal boost and for how many cases they typically use it per month. We also asked how often they incorporate MRI into treatment planning for prostate cancer, how many prostate cancer cases they treat in a typical month, and the degree to which their practice was genitourinary (GU) subspecialized. We asked about fractionation schemes employed when using focal boost, how often radiologists help identify prostate tumors on imaging for treatment planning, barriers to implementing focal boost more often in their practice, and demographic information, including practice setting and years of radiation oncology experience. Finally, we conducted subgroup analyses for respondents from high-income or low-to-middle-income countries, as defined by the World Bank^3^.

## Results

A total of 205 responses were initially collected over a two-week period in December 2022 (12/6/2022-12/20/2022). Due to a low representation of generalists, the survey was then reopened for five days in February 2023 (2/1/2023-2/6/2023) with social media posts requesting more participation from generalists, leading to a total of 263 responses. Those who reported treating zero prostate cancer cases in a typical month were then removed from the study, which lowered the total to 258 responses. The countries and states (for those in the United States) represented by participants are depicted in **Figures 1** and **2**. The highest-represented countries were the United States (42%), Mexico (13%), and the United Kingdom (8%). The majority of respondents (74%) considered their practice to be at least partially GU-focused: 27% completely or nearly completely GU-focused (called hereafter “subspecialists”), 47% partially GU-focused (“partial subspecialists”), and 26% not GU-focused (“generalists”). Additional participant characteristics are provided in **Supplementary Table 1**.

**Figure 1.**
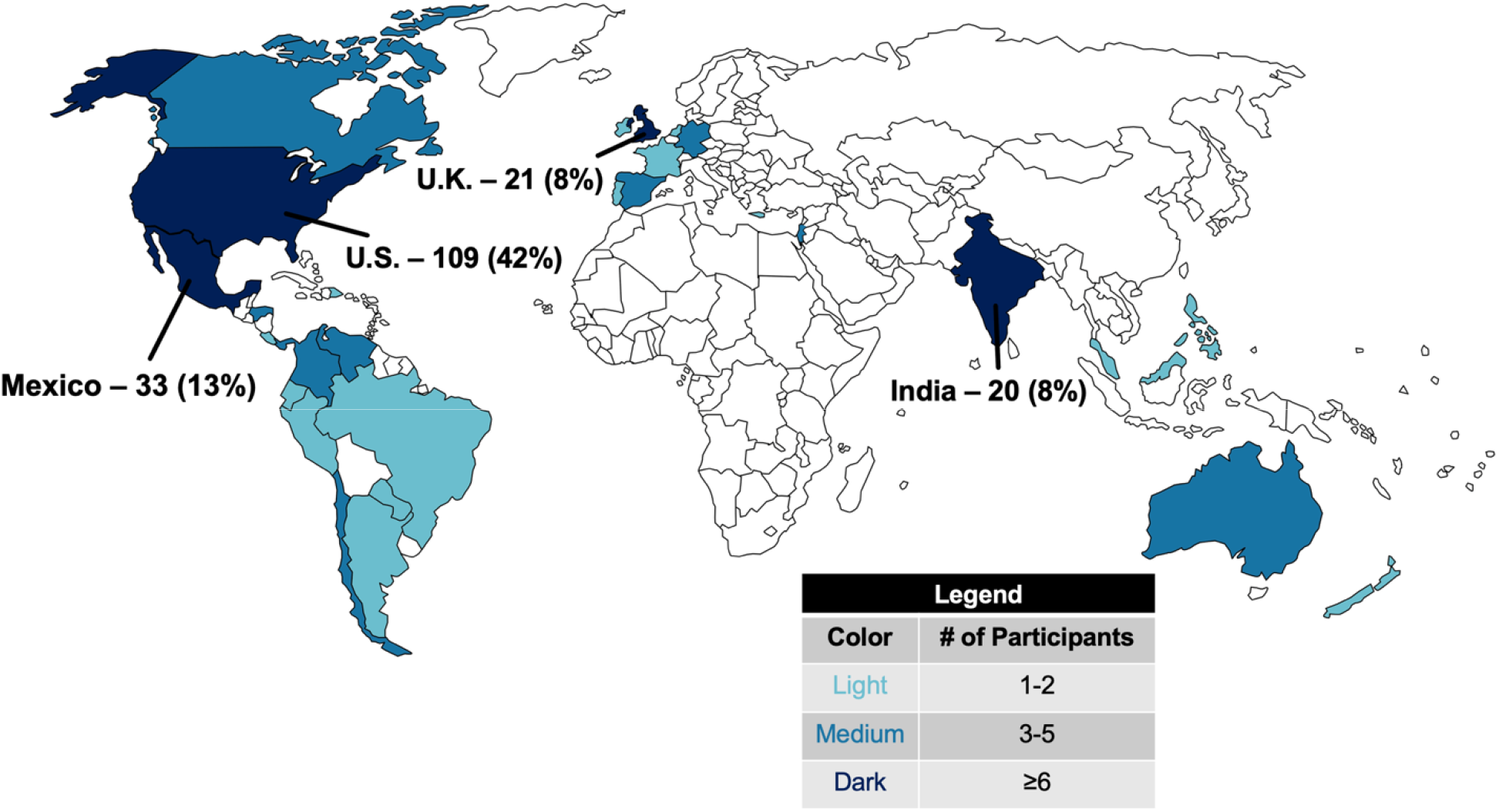
Countries represented by participants.

Overall, 57% of participants do not routinely use focal boost (**Table 1**). Even among complete subspecialists, a substantial proportion (39%) do not routinely use focal boost (**Figure 1**). Less than half of generalists and partial subspecialists, respectively, report routinely using focal boost. Likewise, less than half of participants in both high-income and low-to-middle-income countries routinely use focal boost (**Figure 2**). Additional results are shown in **Table 1**.

**Table 1.** Reported use of intraprostatic focal radiotherapy boost, by category.

| Category | Routinely use focal boost<br>(95% CI) | p-value |
| --- | --- | --- |
| **Degree of GU-subspecialization** |  |  |
| Generalist (n=68) | **31%** (21%, 43%) | 0.0001 |
| Partly subspecialized (n=119) | **40%** (32%, 49%) | 0.002 |
| Completely subspecialized (n=70) | **61%** (50%, 73%) | Reference |
| **Country's income\*** |  |  |
| Low- to middle-income (n=79) | **35%** (25%, 47%) | 0.08 |
| High-income (n=164) | **45%** (37%, 53%) | Reference |
| Declined to state (n=15) | **67%** (40%, 87%) | 0.05 |
| **Practice setting** |  |  |
| Academic medical center (n=133) | **48%** (40%, 56%) | Reference |
| Non-academic hospital (n=21) | **38%** (19%, 62%) | 0.19 |
| Academic-affiliated community hospital<br>(n=38) | **37%** (21%, 53%) | 0.10 |
| Non-academic community hospital<br>(n=20) | **40%** (20%, 60%) | 0.24 |
| Independent/private practice (n=44) | **36%** (23%, 50%) | 0.08 |
| **# of PCa cases treated per month** |  |  |
| 1-4 cases (n=85) | **34%** (25%, 45%) | Reference |
| 5-10 cases (n=97) | **41%** (32%, 52%) | 0.16 |
| >10 cases (n=73) | **55%** (44%, 66%) | 0.004 |

GU: genitourinary; PCa: prostate cancer
\*As defined by the World Bank<sup>3</sup>

**Figure 2.**
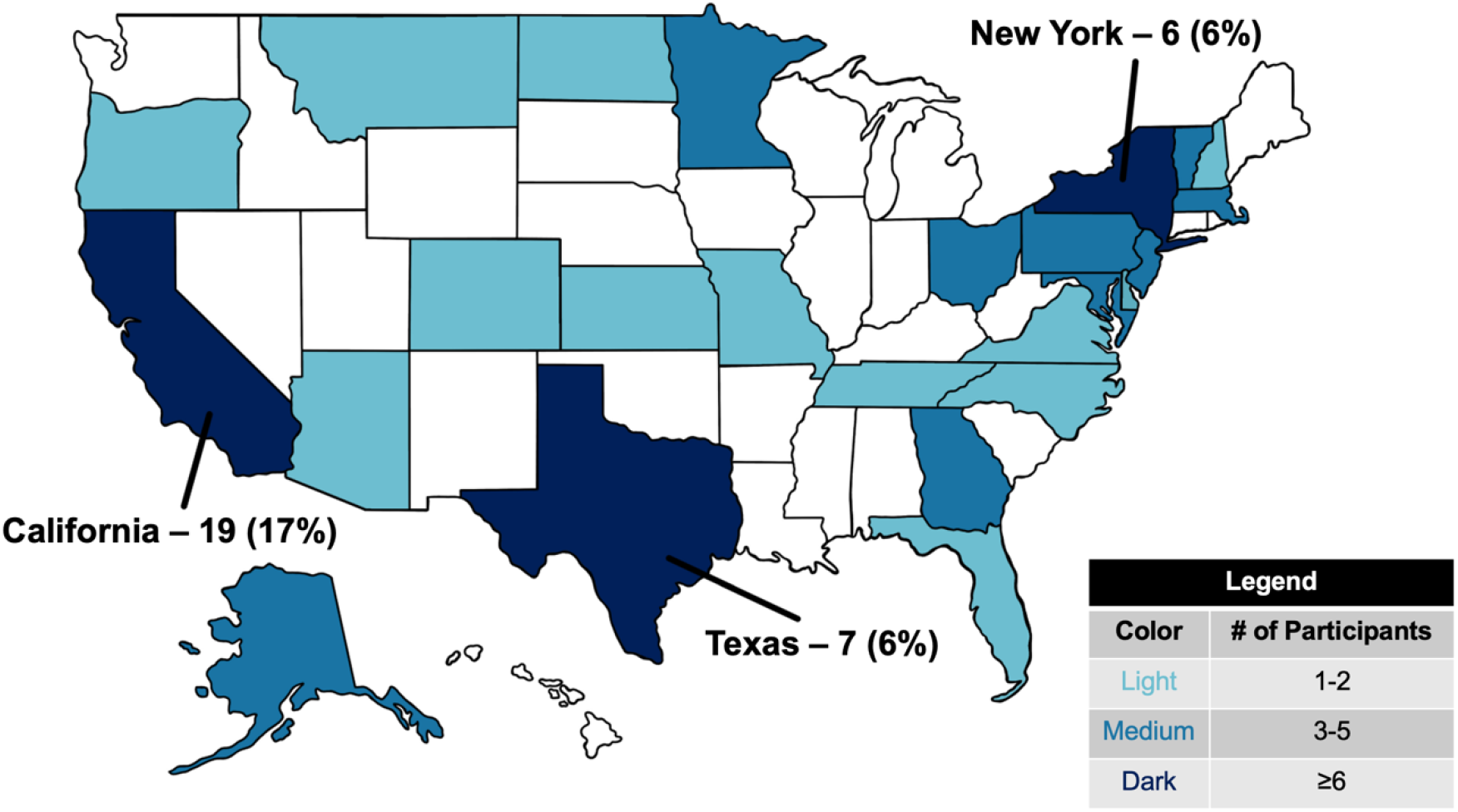
States represented by participants practicing within the United States.

**Figure 3.**
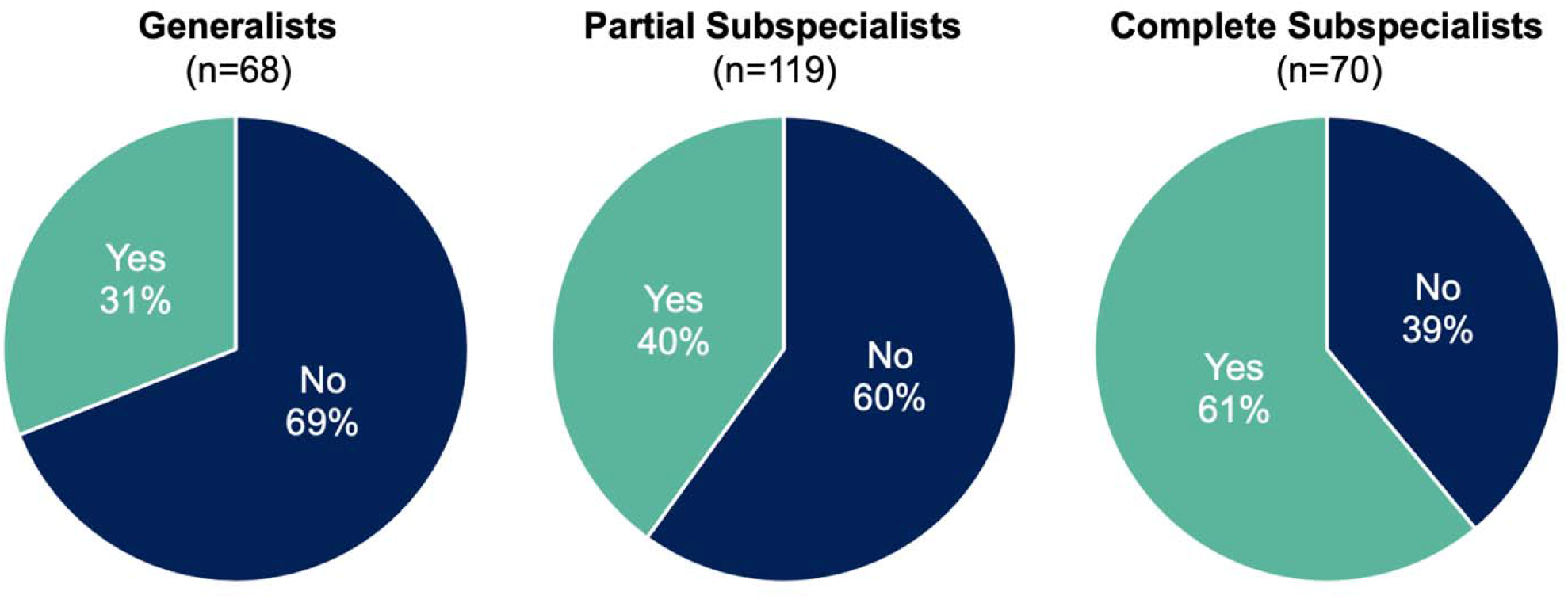
Percentages of participants who routinely use intraprostatic focal boost (“Yes”), by degree of genitourinary (GU)-subspecialization.

**Figure 4.**
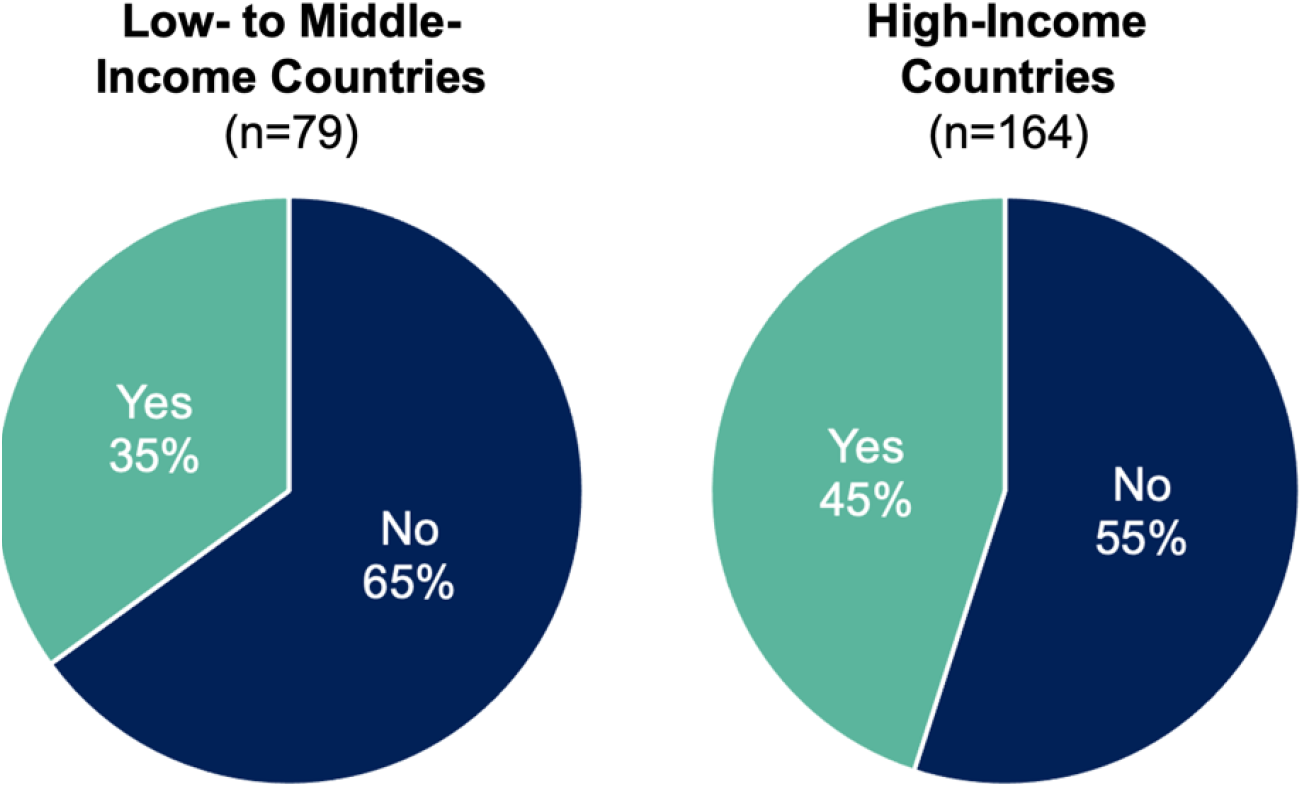
Percentages of participants who routinely use intraprostatic focal boost (“Yes”), by country’s income.

Survey participants’ perceived barriers to implementation are shown in **Table 2**. Write-in answers for other barriers included: not yet part of department protocol; awaiting confirmation of safety and benefit in clinical trials; too large of a tumor or absence of a clear dominant nodule on MRI; lack of standards for lesion delineation; need to justify additional workload of boost planning to physics team; and lack of access to intensity-modulated radiation therapy (IMRT) or intrafraction motion management. Overall, the most commonly cited barriers were concerns about registration accuracy between MRI and CT (37%) and concerns about risk of additional toxicity (35%). Challenges to accessing high-quality MRI were more commonly cited by generalists (32%) and partial subspecialists (34%) compared to complete subspecialists (19%). Generalists more commonly cited lack of training on how to identify prostate tumors on MRI (22%) as a barrier.

**Table 2.**
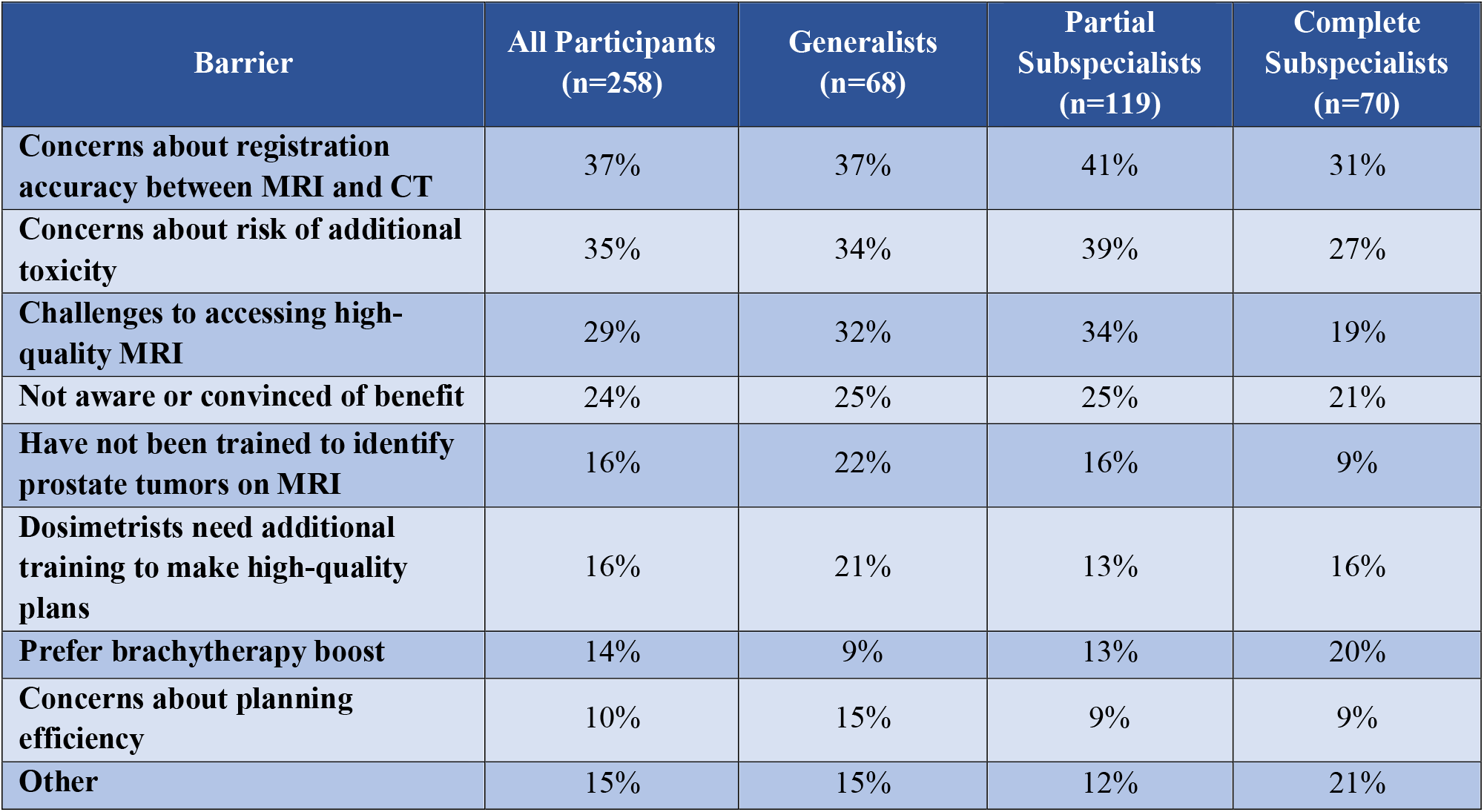
Perceived barriers to implementing intraprostatic focal boost in participants’ respective practices. Participants were asked to select all that apply.

| Barrier | All Participants<br>(n=258) | Generalists<br>(n=68) | Partial<br>Subspecialists<br>(n=119) | Complete<br>Subspecialists<br>(n=70) |
| --- | --- | --- | --- | --- |
| Concerns about registration accuracy between MRI and CT | 37% | 37% | 41% | 31% |
| Concerns about risk of additional toxicity | 35% | 34% | 39% | 27% |
| Challenges to accessing high-quality MRI | 29% | 32% | 34% | 19% |
| Not aware or convinced of benefit | 24% | 25% | 25% | 21% |
| Have not been trained to identify prostate tumors on MRI | 16% | 22% | 16% | 9% |
| Dosimetrists need additional training to make high-quality plans | 16% | 21% | 13% | 16% |
| Prefer brachytherapy boost | 14% | 9% | 13% | 20% |
| Concerns about planning efficiency | 10% | 15% | 9% | 9% |
| Other | 15% | 15% | 12% | 21% |

Among those offering focal RT boost, participants reported using a range of fractionation schemes when including a boost (**Table 3**). The survey question allowed participants to select more than one scheme as applicable to their practice. The most common fractionation scheme overall was moderate hypofractionation (2.1-3 Gy/fraction to the whole prostate) (70%), followed by ultrahypofractionation (≥6 Gy/fraction to the whole prostate) (45%). Generalists appeared to favor using standard fractionation when delivering a focal RT boost more than subspecialists, with 43% of generalists using this scheme compared to 25% of subspecialists. 2% of participants selected “Other” and elaborated that they used brachytherapy boost.

**Table 3.**
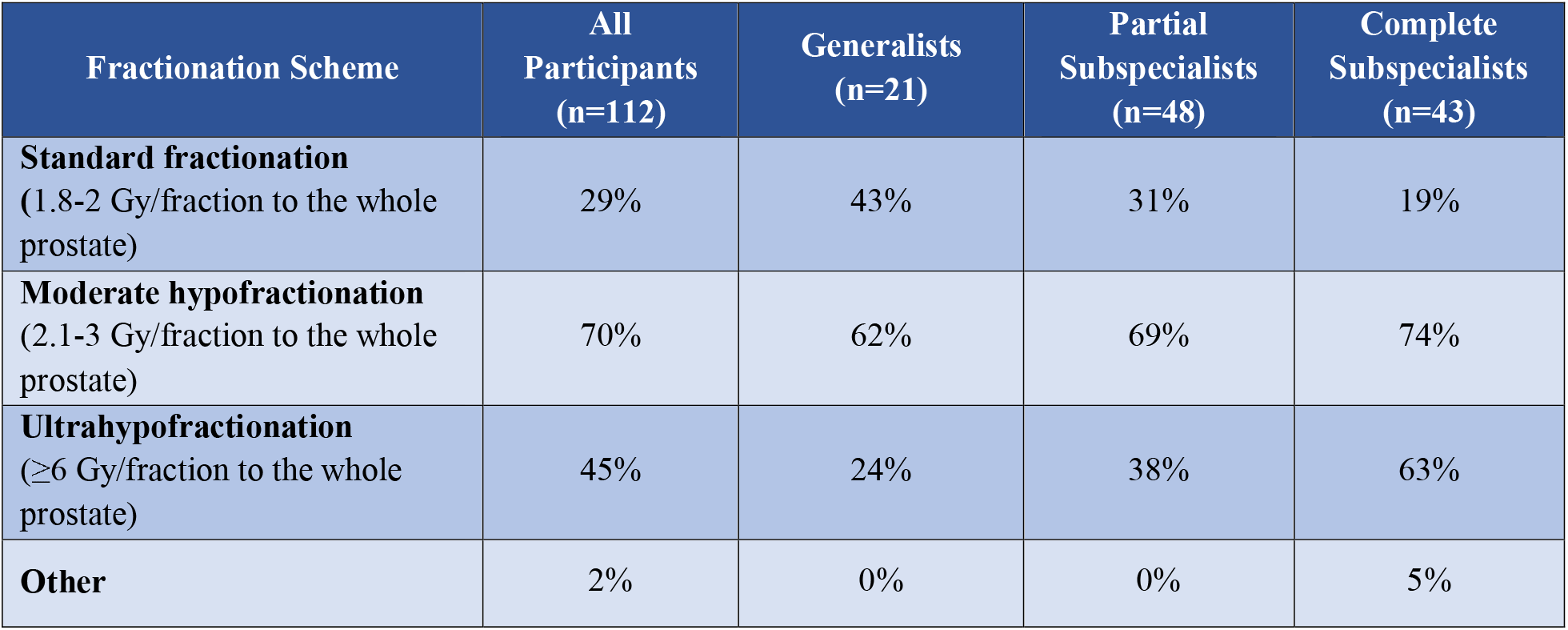
Reported fractionation schemes among participants who routinely use intraprostatic focal boost. Respondents were asked to select all that apply.

| Fractionation Scheme | All Participants (n=112) | Generalists (n=21) | Partial Subspecialists (n=48) | Complete Subspecialists (n=43) |
| --- | --- | --- | --- | --- |
| **Standard fractionation**<br>(1.8-2 Gy/fraction to the whole prostate) | 29% | 43% | 31% | 19% |
| **Moderate hypofractionation**<br>(2.1-3 Gy/fraction to the whole prostate) | 70% | 62% | 69% | 74% |
| **Ultrahypofractionation**<br>($\geq 6$ Gy/fraction to the whole prostate) | 45% | 24% | 38% | 63% |
| **Other** | 2% | 0% | 0% | 5% |

## Discussion

Two years after publication of level 1 evidence supporting focal RT boost, less than half of radiation oncologists responding to our survey have adopted this approach for their patients with prostate cancer. Subspecialists whose clinical practice focuses completely or nearly completely on genitourinary cancers were more likely to report use of focal boost, but a large proportion (39%) of these experts is not routinely using focal boost. Our results show a healthcare disparity exists where only patients seeing certain physicians will even be considered for focal boost.

Participants also provided critical insight into barriers to their increased use of focal boost. Efforts to improve patient outcomes might address the most frequently cited barriers to adoption, including lack of access to high-quality MRI and concerns about accuracy of registration between MRI and CT images. Lack of access to high-quality MRI was more common in low-to-middle-income countries but remained a commonly cited barrier in high-income countries as well. Another commonly perceived barrier among generalists is the lack of training to identify prostate tumors on MRI. Each of these three major barriers could be addressed with improved technology. For example, we have developed a novel prostate cancer MRI biomarker (called the Restriction Spectrum Imaging restriction score, or RSI_rs_) that makes it easier to see clinically significant cancer^4–7^. RSI_rs_ can be obtained on clinical scanners with a 2– 4-minute diffusion-weighted acquisition, in addition to anatomic *T*_*2*_-weighted MRI. In a prospective study, use of RSI_rs_ markedly improved radiation oncologists’ accuracy in identifying prostate tumors^8^.

Some participants expressed doubt about the benefit of focal boost and/or concerns about additional toxicity. The former might be mitigated by inclusion of focal RT boost in continuing education materials and clinical guidelines (it was added to NCCN Guidelines in the past year, specifically for the 35-fraction regimen studied in FLAME^9^). Additional ongoing trials may also corroborate the FLAME results, encouraging adoption. Toxicity concerns are valid. While there was, on average, no increase in toxicity in the focal boost arm in the FLAME trial, focal boost dose on that trial was only escalated to the extent feasible while meeting dose constraints to normal tissues. Additionally, some participants had concerns about focal boost in the setting of larger doses per fraction than used in FLAME. Data on this topic are emerging. Hypo-FLAME was a phase II, single-arm study of ultra-hypofractionation (5 weekly fractions) with focal boost and found acceptable toxicity^10^. Phase III trial evidence is not available for focal boost with ultra-hypofractionated regimens. DELINEATE was a single-center phase II trial in the UK that recently demonstrated safety and feasibility of using focal boost in 20 or 37 fractions. The efficacy and toxicity rates at five years were comparable to those in published trials, including FLAME. PIVOTALboost is an ongoing phase III randomized trial in the UK evaluating focal RT boost in a 20-fraction hypofractionated regimen^11^. Ideal constraints are still under investigation, and some patients may not be good candidates for boosting^12^. On the other hand, if hypofractionation is considered a key barrier to boosting, the logistic advantages of hypofractionation must be weighed against the oncologic benefit of focal boost.

The FLAME trial applied standard clinical techniques in widespread use today. However, additional technologies may play a role in expanding the feasibility of focal RT boost. For example, a posterior tumor may not be amenable to a robust focal tumor boost without violating rectal dose constraints, but placement of a hydrogel spacer could yield more favorable dosimetry for the focal boost. Similarly, adaptive planning and MR-linac platforms could facilitate tighter planning margins and/or more accurate focal boosting. Focal-only brachytherapy boost and intensity modulated proton therapy boost are also possible areas for further study.

Limitations of this study include self-reported practice patterns and a sample of convenience, which led to the overrepresentation of physicians at academic medical centers and of genitourinary subspecialists. The survey was also very brief to encourage participation and does not provide a comprehensive picture of all aspects of practice patterns, including how physicians who do offer focal boost select candidates for this approach or how they identify the target volumes.

In conclusion, from an international survey of over 250 radiation oncologists, we found substantial evidence that most are not routinely offering focal RT boost. This is despite overrepresentation of subspecialists in genitourinary cancers, who might be earlier adopters. Based on commonly cited barriers, adoption of focal RT boost might be accelerated by increased access to high-quality MRI, better registration algorithms of MRI to CT simulation images, more clinical data (especially for larger fraction sizes), physician education on benefit-to-harm ratio, and physician training on how to contour prostate lesions on MRI. Addressing these barriers would likely increase the adoption of focal RT boost and improve the efficacy of RT for more patients with prostate cancer.

## Data Availability

All data produced in the present study are available upon reasonable request to the authors.

## Acknowledgments

Dr. Tree acknowledges the support of Cancer Research UK (grant numbers C7224/A28724 and C33589/A28284) as well as NHS funding to the NIHR Biomedical Research Centre at The Royal Marsden and The Institute of Cancer Research. The views expressed in this publication are those of the author(s) and not necessarily those of the NHS, the National Institute for Health Research or the Department of Health and Social Care. Dr. Lui acknowledges the support of the Grillo-Marxuach Family Fellowship to the Moores Cancer Center at UC San Diego. Drs. Lui and Seibert also acknowledge support from the Radiological Society of North America. Dr. Seibert acknowledges the support of the National Institutes of Health (grant numbers NIH/NIBIB K08EB026503, and NIH UL1TR000100), the American Society for Radiation Oncology, and the Prostate Cancer Foundation.

---

**Supplementary Table 1.**
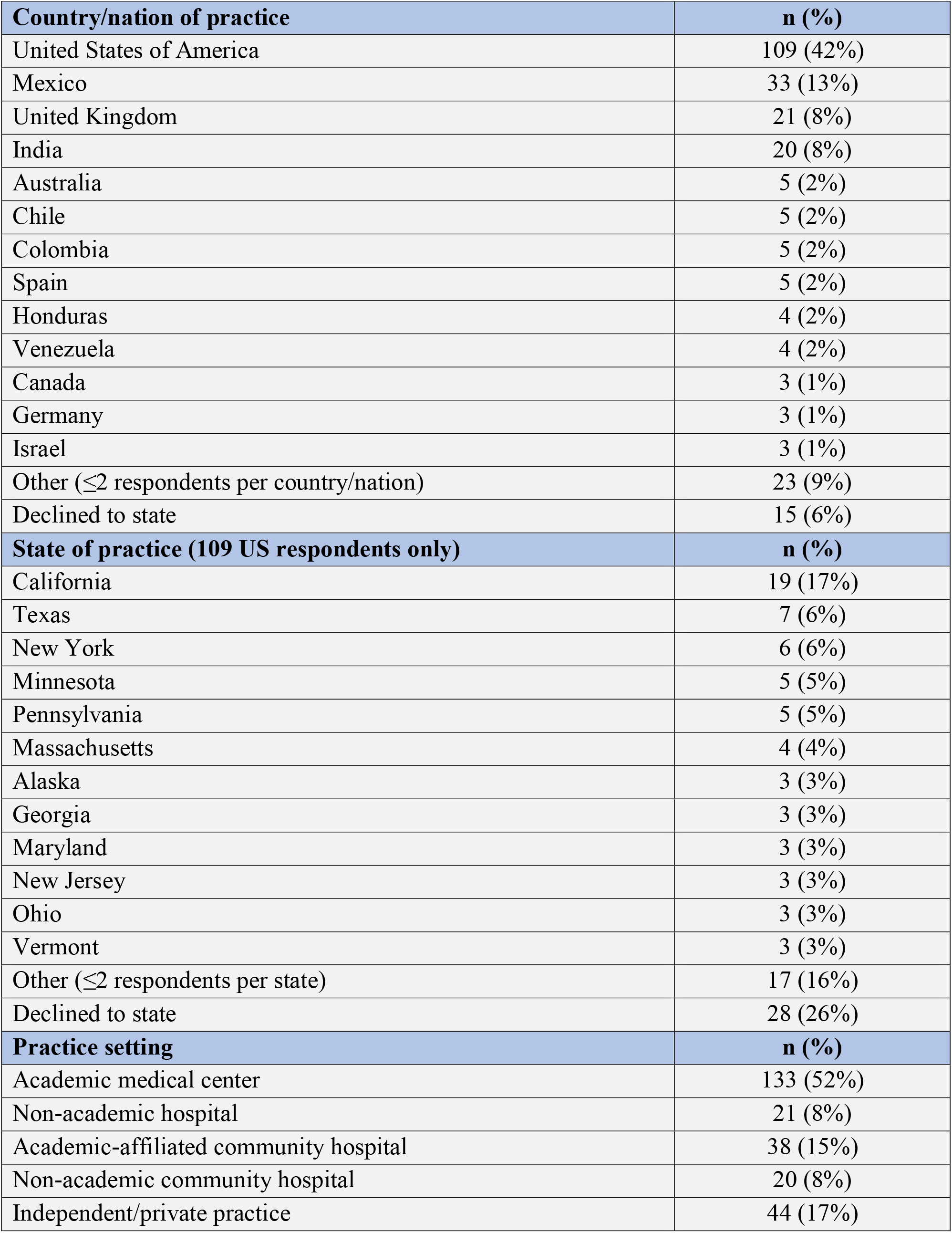
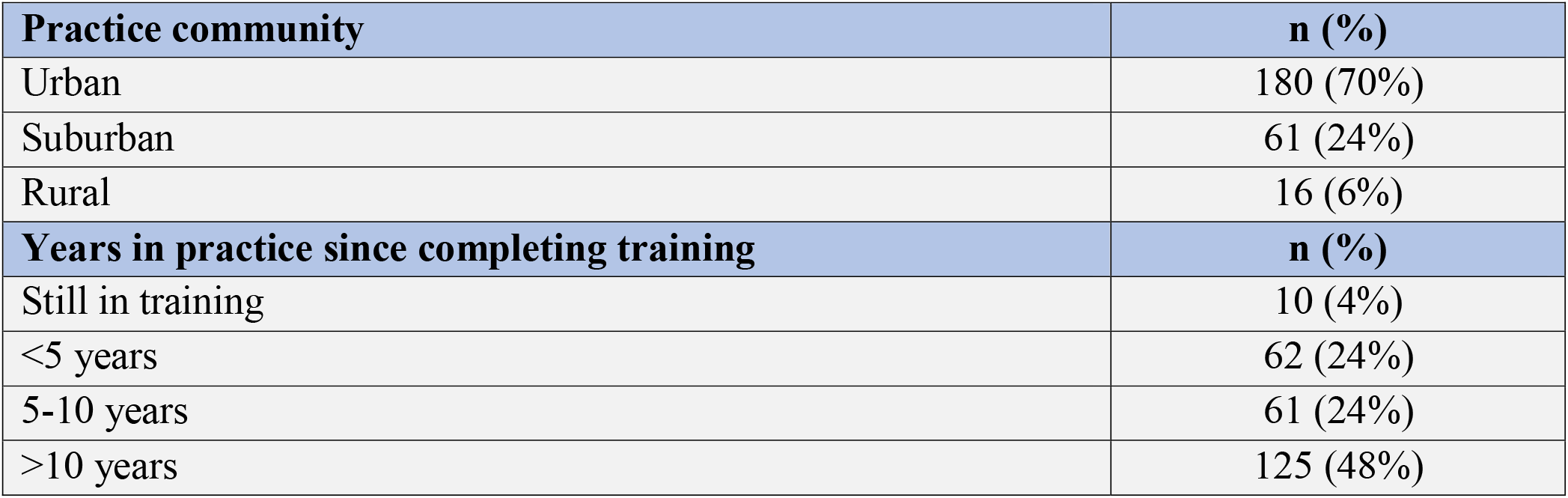
Participant characteristics.

