## Supplementary Material: Survey Questions for "Use of focal radiotherapy boost for prostate cancer and perceived barriers toward its implementation: a survey" (from another version of this preprint: version 1, 2023-02-02)

**Prostate Focal Boost Survey**

1. Do you consider your practice genitourinary (GU) focused/subspecialized?
   A. No
   B. Yes, partially focused on GU
   C. Yes, completely or nearly completely focused on GU
2. How often do you incorporate MRI into your treatment planning for intact prostate cancer?
   A. Never
   B. <1/3 of cases
   C. 1/3 to 2/3 of cases
   D. >2/3 of cases
3. How many intact prostate cancer cases do you typically contour per month?
   A. None
   B. 1-4 cases
   C. 5-10 cases
   D. >10 cases
4. How many cases per month (typically) do you use a tumor-directed boost within the prostate (i.e., to the dominant intraprostatic lesion or visible tumor)?
   A. None
   B. 1-4 cases
   C. 5-10 cases
   D. >10 cases
5. (Skip if answer to #4 is “None”) When you use a tumor-directed boost, how often has a radiologist directly helped identify the tumor for you on the images (i.e., not just with a description in the report)? Examples include arrows pointing to the lesion, contouring the lesion, or reviewing your contours for accuracy.
   A. Never
   B. <1/3 of cases
   C. 1/3 to 2/3 of cases
   D. >2/3 of cases
6. (Skip if answer to #4 is “None”) Which fractionation schemes (i.e., how many total fractions in the radiotherapy course) do you use when including a tumor-directed boost? (Check all that apply)
   A. Standard fractionation (1.8-2 Gy/fraction to the whole prostate)
   B. Moderate hypofractionation (2.1-3 Gy/fraction to the whole prostate)
   C. Ultrahypofractionation (≥6 Gy/fraction to the whole prostate)
   D. Other: [Text]
7. In your practice, what are the important barriers to using tumor-directed boost more often? (Check all that apply)
   A. Not aware of or convinced about benefit
   B. Concerns about risk of additional toxicity
   C. Challenges to accessing high-quality MRI
   D. Concerns about registration accuracy between MRI and CT
   E. Have not been trained to identify prostate tumors on MRI
   F. Dosimetrists need additional training to make high-quality plans
   G. Concerns about planning efficiency
   H. Prefer brachytherapy boost
   I. Other: [Text]

Demographic Information

8. What country do you work in? [Drop-down menu]
9. (If answer to #8 is “United States”, otherwise skip) What state do you work in? [Drop-down menu]
10. Which of the following best describes your practice setting?
    A. Academic medical center
    B. Non-academic hospital
    C. Academic-affiliated community hospital
    D. Non-academic community hospital
    E. Independent/private practice
    F. Other: [Text]
11. Which of the following best describes the community in which you work?
    A. Urban
    B. Suburban
    C. Rural
12. Since completing your clinical training, how many years have you been in clinical practice?
    A. Still in training
    B. <4 years
    C. 5-10 years
    D. >10 years
